# A Light-weight Text Summarizer for Fast Access to Medical Evidence

**DOI:** 10.1101/2020.05.22.20110742

**Authors:** Abeed Sarker, Yuan-Chi Yang, Mohammed Ali Al-Garadi

## Abstract

The performances of current medical text summarization systems rely on resource-heavy domain-specific knowledge sources, and preprocessing methods (e.g., classification or deep learning) for deriving semantic information. Consequently, these systems are often difficult to customize, extend or deploy in low-resource settings, and are operationally slow. We propose a fast summarization system that can aid practitioners at point-of-care, and, thus, improve evidence-based healthcare. At runtime, our system utilizes similarity measurements derived from pre-trained domain-specific word embeddings in addition to simple features, rather than clunky knowledge bases and resource-heavy preprocessing. Automatic evaluation on a public dataset for evidence-based medicine shows that our system’s performance, despite the simple implementation, is statistically comparable with the state-of-the-art.

## Introduction

Current clinical practice guidelines urge practitioners to follow the principles of evidence-based medicine, which requires them to integrate the best external scientific evidence with clinical expertise.^1,2^ Early and recent studies show that one of the biggest obstacles to evidence-based medicine practice is information overload caused by the large volume of medical literature available.^3^ Searching through medical evidence is too time consuming and practitioners often consider the task to be unproductive.^4^ A standard clinical query on PubMed,^1^ which indexes over 30 million articles, typically returns multiple pages of research publications. Hersh *et al*.^5^ discussed that it takes more than 30 minutes for a practitioner to search for an evidence-based answer, and, particularly at point-of-care, practitioners cannot afford to spend that much time. Literature searching, and fast access to relevant information, is particularly beneficial for medical students and young practitioners because of their lack of clinical experience.

Motivated by the importance of incorporating scientific evidence in everyday medicine practice, automated text summarization research has attempted to address the problem. In particular, query-focused text summarization approaches have been explored to aid evidence-based medicine.^6–8^ These systems take queries (in natural language or key-terms) as input and output query-relevant summaries. In terms of automatic summary qualities, the performances of successful approaches designed for the medical domain have relied heavily on domain-specific knowledge sources.^9^ For example, one study Demner-Fushman and Lin^10^ incorporated sentence-level knowledge in a supervised classification system trained to detect *outcome* sentences. Sarker *et al*.^8^ and ShafieiBavani^7^ utilized manually annotated summarization datasets specific to the domain to generate extractive and abstractive summaries—both systems relying heavily on the identification of domain-specific generalizations, concepts, and associations. Hristovski *et al*.^11^ proposed the use of domain-specific semantic relations for performing question answering for biomedical literature. Further discussion of such systems is outside the scope of this short paper, and detailed descriptions of medical text summarization systems over the years are available through survey papers.^12,13^

Adaptation of summarization systems to a particular domain can be computationally expensive, and require large numbers of external tools.^14^ Within the medical domain, systems typically use the Unified Medical Language System,^2^ and tools such as MetaMap^15^ are employed to derive domain-specific knowledge from lexical representations. This is in turn used in downstream tasks, or as features in learning systems. Heavy dependence on these domain-specific resources introduces disadvantages: (i) the systems are not very portable, (ii) they are difficult to re-implement and/or deploy without the knowledge sources, and (iii) the systems are slow and resource-heavy.

The goal of our work is to design a light-weight and fast medical text summarization system that is decoupled from knowledge bases. In our view, such a system may be operationalized as a web-based service, enabling practitioners to review medical evidence at point-of-care. The proposed system relies on publicly available labeled and unlabeled data, dense word vectors learned from the unlabeled data, and a set of simple features that require little computational resources and time. Automatic evaluation of our system with a state-of-the-art system on a standard dataset shows that our approach produces comparable summary quality.

## Methods

We use a public dataset—the corpus provided by Molla-Aliod *et al*.,^16^ which contains a total of 456 questions along with expert-authored single- and multi-document evidence-based summarized responses to them. Each question is generally associated with multiple single-document summaries, which present evidence from distinct studies. The abstracts of the studies from which the answers were derived are made available from PubMed. In total, the corpus contains 2,707 single-document summaries. To ensure fair comparison, we obtained the exact train-test split from the authors of the QSpec system^8^ (1,388 for training, 1,319 for evaluation). The training set is used to devise feature scoring techniques and learn weights for all the feature scores. The summary sentences are scored as the sum of the weighted feature scores, and chosen sequentially (from first to last), taking into consideration the target sentence position and the contents of the chosen sentences. The scoring process can be summarized as: 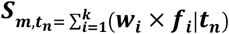 where 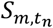 is the score for sentence number *m* of a text, given the summary target sentence number *t_n_*, and *w_i_* and *f_i_* are the weight and score for feature *i*, respectively. For each *t_n_*, the top-scoring sentence is chosen. Following experimentation with multiple features, we selected five that could be computed fast, and proved to be useful when used in combination. We describe these in the following paragraphs.

### Word Embedding-based Maximal Marginal Relevance

Maximal Marginal Relevance (MMR)^17^ is a strategy that can be used to increase relevance and reduce redundancy in summarization. The core of the technique relies on computing two similarity measures—between sentences and the associated query, and between the sentences themselves. During score generation, sentences are rewarded for being similar to the query, while at the same time, they are penalized for being similar to sentences that have already been chosen to be included in the summary. The similarity values are combined linearly with suitable weights 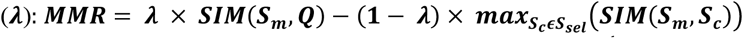, where ***SIM*(*S_m_,Q*)** is the similarity score between a sentence and the question and 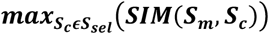 is the maximum similarity between the same sentence and the set of already chosen summary sentences. Choosing the best three-sentence summary is a combinatorial optimization problem, and MMR enables us to approach sentence selection in a sequential manner.

We experimented with two variants of MMR that rely on the distributed representations of the words in the sentences and the questions. We obtained pre-trained embeddings that were generated from all PubMed and PMC OA texts^18^ using the *word2vec* tool^3^ (vector = 200, window size = 5) and the *skip-gram* model.^19^ For the first variant, we compute the similarity between two text segments (*i.e*., sentence vs. question and sentence vs. sentence) as the *average* cosine similarity of all the terms. We compute this average by adding the cosine similarities of all the term combinations and dividing by the product of the lengths of the two texts. For the second variant, we use the word vectors in a text segment to compute its centroid in vector space. A single centroid is computed for the set of all words within the set of already chosen sentences **(*S_sel_*)**. These centroids are then used to compute MMR.

### Traditional MMR Score

For the traditional MMR score, we first preprocessed the terms by lowercasing, stemming and removing stop words. We then computed the *tf × isf* for each word in a sentence and the question**—**where *tf* is the frequency of a term in a text segment and *isf* is the *inverse sentence frequency* of the term in all the texts (*i.e*., the inverse of how many sentences including the question contain the term). We then generate vectors for each sentence using the *tf × isf* values of the terms.

### Sentence Length Score

Sentence length is a metric that may filter out uninformative, short sentences by assigning them a lower score, while rewarding sentences that are relatively longer in a document. In past research in this domain, it has been mentioned to be particularly effective for tie-breaking. In our work, we attempted to assign penalties to very short sentences (e.g., 1—3 word sentences), which often represent section headers. At the same time, our goal was to assign higher scores to longer sentences---with decreasing gradients for very long sentences, such that this score does not play a significant role in choosing between those informative sentences.

Our experiments on the training set suggested that a *sin* function conveniently served this purpose. The average sentence length in the training data is approximately 150 characters, so we considered 0 and 300 characters to be the lower and upper length limits, respectively, and mapped the lengths to the range [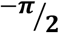, 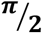]. Following that, we applied a *sin* function to the mapped value to generate a length score between [-1,1] (Figure 1).

**Figure 1.**
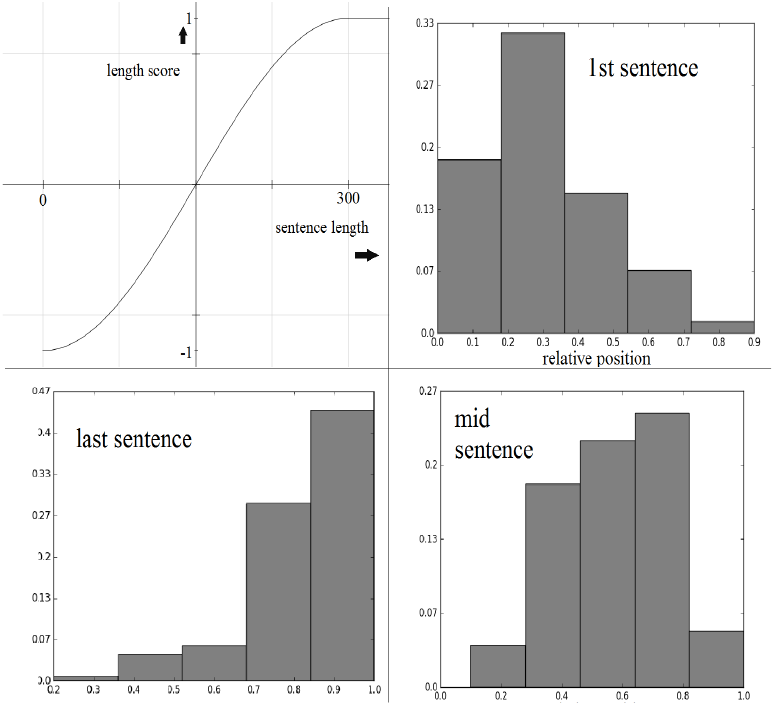
Clockwise from top-left: sine function for sentence length score, maxed at 300 characters; first, middle and last sentence relative position distributions from the best-scoring extractive training set summaries.

### Sentence Position Score

Our last score is based on sentence position and the target sentence number. Sentence position has been shown to be a crucial metric for extractive summarization in domains including news^20^ and medical.^10^ We use an identical scoring approach to Sarker *et al*.^8^ and apply target sentence specific summarization. We first obtain the best three-sentence summary for each training text, and use these sentences to generate normalized frequency distributions of the relative sentence positions for these *best* sentences for each of the three target sentence positions. During summary generation, given the relative sentence position *r* of a source sentence, the score assigned is the normalized frequency for *r* in the given target sentence distribution.

### Weight Optimization and Sentence Scoring

We compute optimal weights for scoring using the training set via a grid search in the range [0.0,1.0] with step sizes of 0.1. For each weight combination, all the three-sentence training set summaries are generated and the ROUGE^21^ summary evaluation tool is used to compare the extractive summaries with the expert-authored summaries in the corpus. The weights producing the highest F_1_-score for the training set are used for evaluation on the test set.

## Evaluation, Results and Discussion

As mentioned, we used the ROUGE summary evaluation tool to compare the performance of our system with other systems. Since the summaries are not length-restricted, we used the ROUGE-L F_1_-score as our evaluation metric. ROUGE scores have been shown to be highly correlated with human evaluators.^21^ Table 1 presents the performance of our system along with several other systems, including the state-of-the-art for this task (QSpec). Identical training-test splits were used for evaluation. The table shows that despite the simplicity of our approach, its performance is comparable to the state-of-the-art, and significantly better than other baselines.

**Table 1.** Comparison of ROUGE-L F_1_-scores for our summarizer with other systems and 95% confidence intervals.

| System | ROUGE-L F<sub>1</sub> Score | 95% CI |
| --- | --- | --- |
| Our system | 0.166 | 0.162 – 0.170 |
| Sarker et al.<sup>8</sup> | 0.168 | 0.164 – 0.172 |
| Last 3 Sentences | 0.155 | 0.151 – 0.158 |
| Demner-Fushman and Lin<sup>10</sup> | 0.159 | 0.152 – 0.164 |
| Random | 0.154 | 0.150 – 0.157 |
| First 3 Sentences | 0.140 | 0.136 – 0.143 |

Due to the simplicity of our approach, it can be easily re-implemented, customized or extended for real-life settings, and the results can be reproduced without requiring the use of third-party tools. Compared to the resource-heavy QSpec system, which requires query and sentence classification, and the generation of UMLS semantic types and associations, our approach requires minimal preprocessing. Only a set of pre-trained word embeddings are required. The light-weight nature of the summarizer also means that it runs faster than QSpec. On a standard computer (Intel® i5 2.0 GHz processor), it takes our summarizer a few minutes to summarize all the documents in the test set.

We obtained the word embeddings from past research and used them without modification. There is a possibility that the learning of these embeddings can be customized to the summarization task for improving performance. Furthermore, since the embeddings can be learned from publicly available data, and no other resources are required by the summarizer, the system can be implemented anywhere with minimal effort.

## Conclusions and Future Work

In this paper we presented a simple, query-focused, extractive summarization system suited for application in the domain of evidence-based medicine. Unlike past systems in this domain that rely heavily on domain-specific resources such as knowledge bases, our system is light-weight in nature and relies only on distributed representations of words learned from unlabeled text. Using a set of similarity-based and structural features, our system performs comparably to the state-of-the-art system, with a ROUGE-L F_1_-score of 0.166. Because of the simplicity of our system, it can easily be ported and implemented in different settings.

## Data Availability

Corpus used is publicly available.

https://sourceforge.net/projects/ebmsumcorpus/

## Footnotes

1 https://www.ncbi.nlm.nih.gov/pubmed/ [Access date: 25th Nov 2019].

2 http://www.nlm.nih.gov/research/umls/ [Access date: 12th Dec 2019].

3 https://code.google.com/p/word2vec/. [Access date: 17th Nov 2019].

